# Prevalence of Chemosensory Dysfunction in COVID-19 Patients: A Systematic Review and Meta-analysis Reveals Significant Ethnic Differences

**DOI:** 10.1101/2020.06.15.20132134

**Authors:** Christopher S. von Bartheld, Molly M. Hagen, Rafal Butowt

**Affiliations:** Center of Biomedical Research Excellence in Cell Biology, University of Nevada, Reno School of Medicine, Reno, Nevada, USA; School of Community Health Sciences, University of Nevada, Reno, Nevada, USA; Department of Molecular Cell Genetics, L. Rydygier Collegium Medicum, Nicolaus Copernicus University, Bydgoszcz, Poland

**Keywords:** Anosmia, Smell, COVID-19, SARS-CoV-2, Prevalence, Diagnosis, Hypogeusia, Taste, Ethnicity

## Abstract

A significant fraction of people who test positive for COVID-19 have chemosensory deficits. However, the reported prevalence of these deficits in smell and/or taste varies widely, and the reason for the differences between studies is unclear. We determined the pooled prevalence of such chemosensory deficits in a systematic review. We searched the COVID-19 portfolio of the National Institutes of Health for all studies that reported the prevalence of smell and/or taste deficits in patients diagnosed with COVID-19. Forty-two studies reporting on 23,353 patients qualified and were subjected to a systematic review and meta-analysis. Estimated random prevalence of olfactory dysfunction was 38.5%, of taste dysfunction was 30.4% and of overall chemosensory dysfunction was 50.2%. We examined the effects of age, disease severity, and ethnicity on chemosensory dysfunction. The effect of age did not reach significance, but anosmia/hypogeusia decreased with disease severity, and ethnicity was highly significant: Caucasians had a 3-6 times higher prevalence of chemosensory deficits than East Asians. The finding of ethnic differences points to genetic, ethnicity-specific differences of the virus-binding entry proteins in the olfactory epithelium and taste buds as the most likely explanation, with major implications for infectivity, diagnosis and management of the COVID-19 pandemic.

## Introduction

The first reports of disturbances of smell and taste in COVID-19 patients emerged in February and March of 2020. Initially, these reports were anecdotal, but soon articles consistently described an increased prevalence of chemosensory deficits. Many of the earliest studies were compiled in six recent reviews (da Costa et al., 2020; Passarelli et al., 2020; Pellegrino et al., 2020; Printza and Constantinidis, 2020; Sedaghat et al., 2020; Tong et al., 2020). Two of these reviews (Passarelli et al., 2020; Tong et al., 2020) conducted a meta-analysis of 6 and 10 studies, respectively, the other four were narrative reviews (da Costa et al., 2020; Pellegrino et al., 2020; Printza and Constantinidis, 2020; Sedaghat et al., 2020). Studies reported prevalence of chemosensory dysfunction with wide ranges, between 5% and 98% for anosmia, and between 6% and 93% for taste dysfunctions (Tong et al., 2020). The reasons for differences in the prevalence reported in different studies were thought to be due to differences in the age of patients, in assessment methods, or in the severity of the disease. In addition, patient selection was thought to play a role – since some data were from hospitalized patients, others from clinic visits, and cohorts were from different countries, and data obtained with different study designs. Most studies relied on the patients’ subjective impressions about sensation of smell or taste.

To gain a more comprehensive and conclusive account of the prevalence of chemosensory deficits in COVID-19, we conducted a systematic review and meta-analysis of 42 studies that reported on the chemosensory function of 23,353 patients diagnosed with COVID-19. We included preprints of not yet peer-reviewed studies, up to the posting on June 10, 2020. Because we considered a larger number of studies and larger cohort numbers than previous reviews, we provide a clearer picture of the true prevalence, and, importantly, we stratified and examined confounding variables such as age, methodology, disease severity, and ethnicity. We report that ethnicity is a significant factor. The finding of ethnic differences has important implications for the diagnosis of COVID-19, and for the management of the pandemic in countries with different ethnic populations.

## Results

We adhered to the preferred reporting items for systematic reviews and meta-analyses (PRISMA), as shown in the flowchart (Fig. 1). Our search strategy retrieved 42 studies that fulfilled the inclusion criteria, reporting on 46 distinct cohorts, with prevalence information on a total of 23,353 patients from 18 different countries. Studies are from multiple countries (Lechien et al., 2020a; Qiu et al., 2020), from Italy (Giacomelli et al., 2020; Vaira et al., 2020b; Spinato et al., 2020; Vaira et al., 2020c; Gelardi et al., 2020; Meini et al., 2020; Boscolo-Rizzo et al., 2020), Germany (Streeck, 2020; Luers et al., 2020; Bertlich et al., 2020; Hornuss et al., 2020; Haehner et al., 2020; Qiu et al., 2020), France (Klopfenstein et al., 2020; Lechien et al., 2020b; Zayet et al., 2020; Tudrej et al., 2020; Qiu et al., 2020), Spain (Beltran-Corbellini et al., 2020; Borobia et al., 2020; Abalo-Lojo et al., 2020; Romero-Sanchez et al., 2020), USA (Yan et al., 2020a; Yan et al., 2020b; Kaye et al., 2020; Menni et al., 2020), China (Mao et al., 2020; Qiu et al., 2020), Korea (Rabin, 2020; Lee et al., 2020), Singapore (Wee et al., 2020; Kai Chua et al., 2020), UK (Patel et al., 2020; Menni et al., 2020), Canada (Carignan et al., 2020; Lee et al., 2020), Iceland (Gudbjartsson et al., 2020), Israel (Levinson et al., 2020), Iran (Moein et al. 2020), Holland (Tostmann et al., 2020), Switzerland (Speth et al., 2020), Belgium (Lechien et al., 2020c), Greece (Tsivgoulis et al., 2020), and Japan (Komagamine and Yabuki, 2020). The included studies are listed chronologically and by geographic region (East Asia vs Europe/ Middle East/ North America) in Table 1. Whether some patients had both, smell and taste dysfunction was stated explicitly only in a fraction of studies or was apparent from the numbers given (19/46 cohorts, Table 1).

**TABLE 1.**
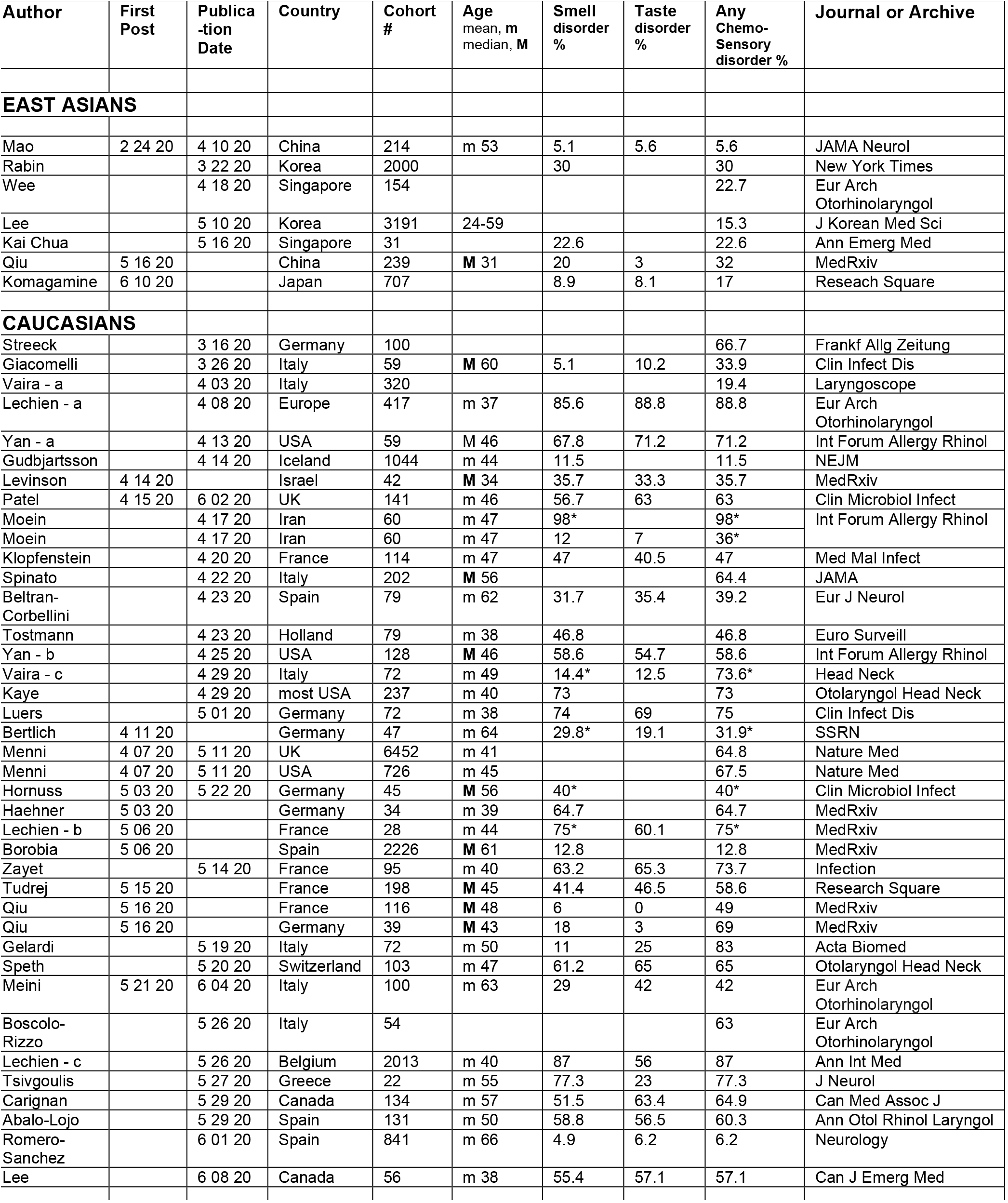
Smell and Taste Dysfunction in COVID-19: Pooled Analysis - Chronology of Studies.

**Figure 1.**
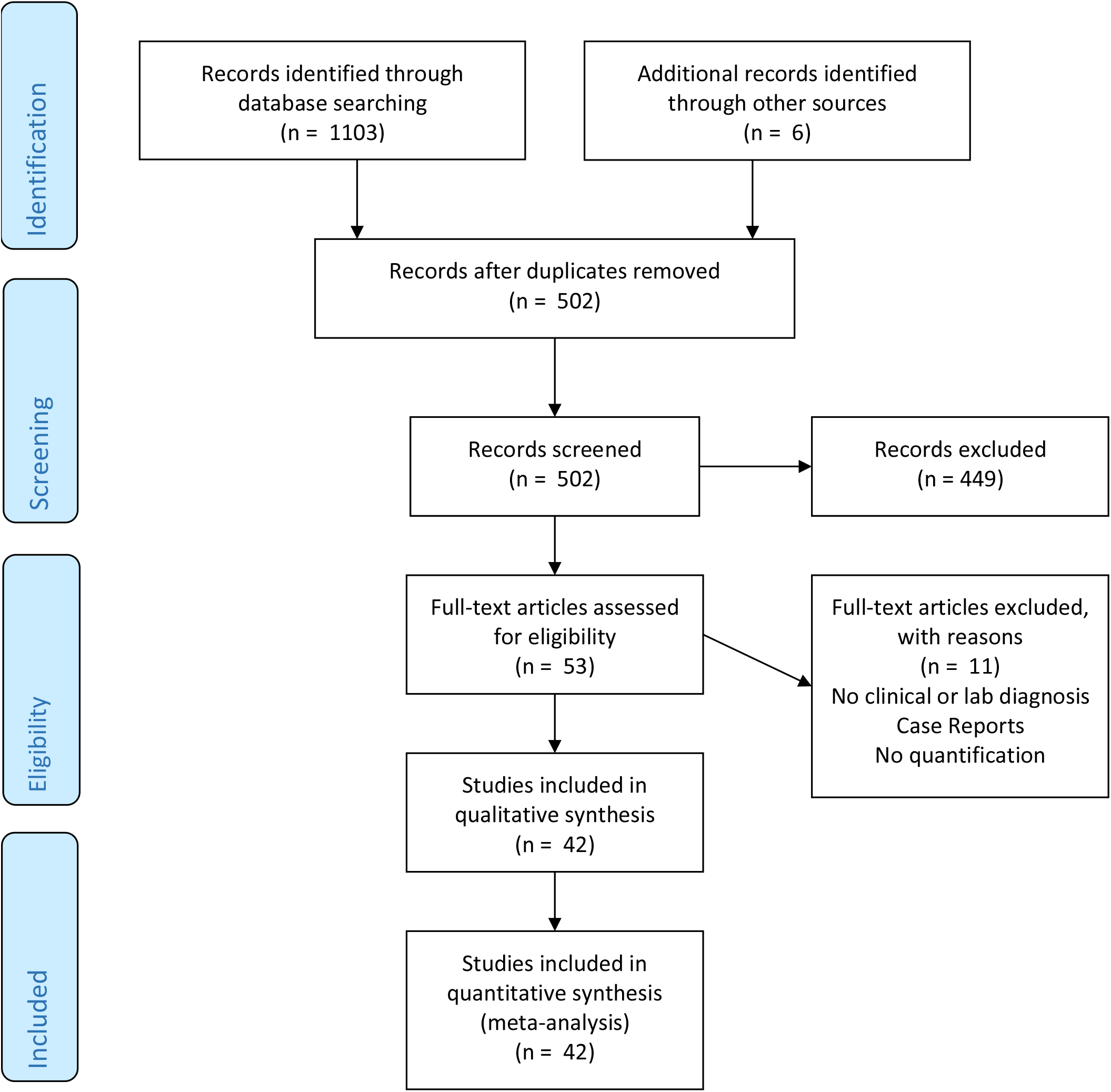

The overall estimated random prevalence of smell loss among COVID-19 patients, calculated from a total of 38 cohorts containing 12,154 persons, was 38.48% [95% confidence interval (CI), 28.33-49.74%]. The meta-analysis indicated that between-study variability in prevalence of smell loss was high (τ^2^ = 1.99; heterogeneity I^2^ = 98.9% with Q = 3363.4, *df* = 37 and *p* = 0.999; Higgins and Thompson, 2002) and examination of the funnel plots, as expected, showed evidence of publication bias (Fig. 2A). The 30 cohorts with information on taste loss contained a total of 9,589 patients. The overall estimated random prevalence of taste loss among COVID-19 patients was 30.37% [95% CI, 20.07-43.11%]; the analysis indicated that between-study variability was high (τ^2^ = 2.2449; heterogeneity I^2^ = 98.8% with Q = 2341.1, *df* = 29 and *p* = 0.999; Fig. 2B). When smell and taste loss were combined, the overall estimated random prevalence obtained from 23,353 patients in 46 cohorts was 50.20% [95% CI, 41.51-58.88%]; the analysis showed high heterogeneity with some publication bias (Fig. 2C).

**Figure 2.**
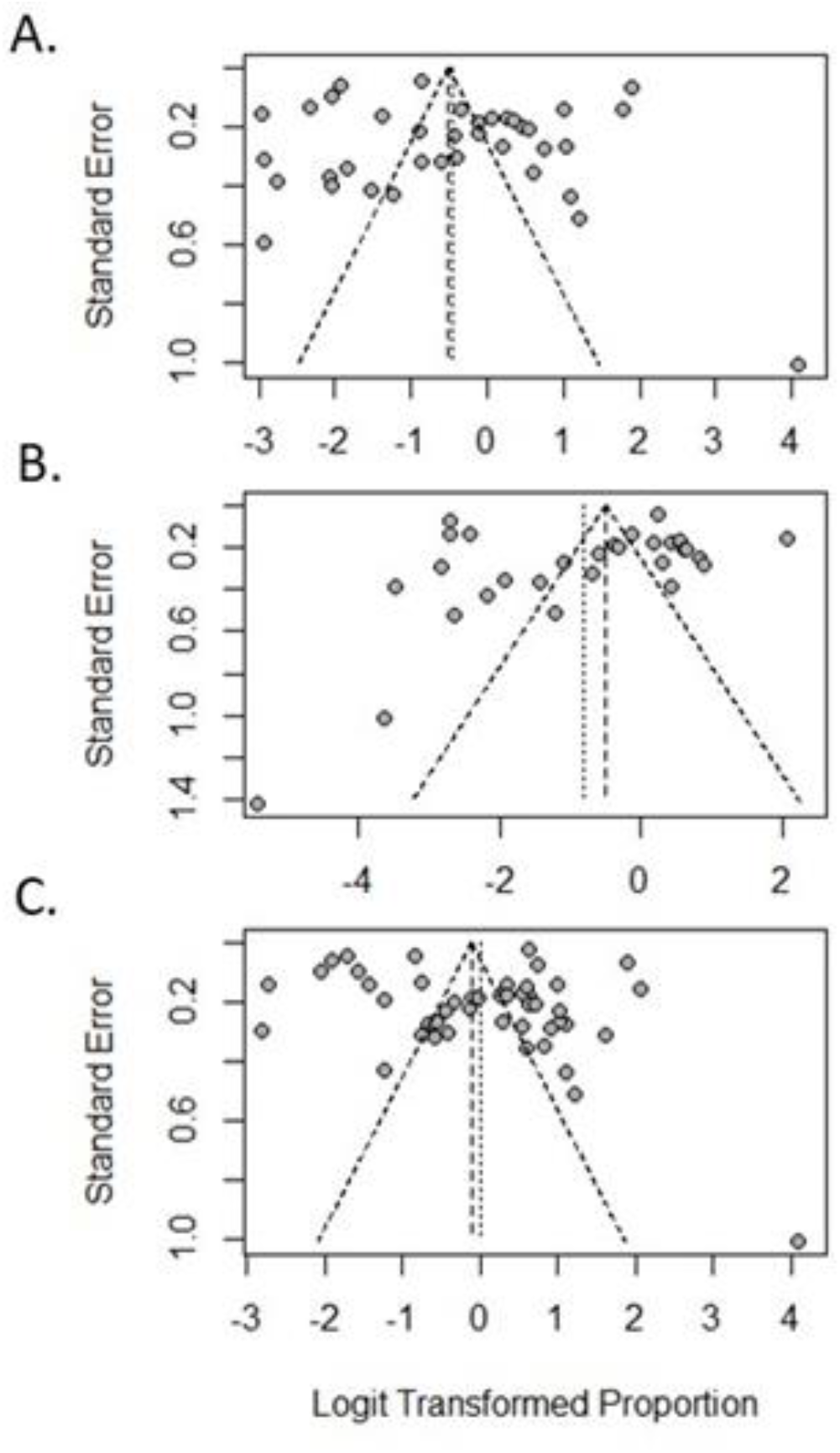
Funnel Plots of the prevalence of dysfunction of smell (A), taste (B), and smell and/or taste (C) in COVID-19 patients. Each dot represents a single study with the x-axis showing the logit transformed proportion of people in each study that lost their sense of (A) smell, (B) taste, and (C) smell and/or taste; the y-axis shows the standard error.

There was a significant difference between countries with majority East Asians and countries with majority Caucasians in the prevalence of smell, taste and any chemosensory dysfunction. Ethnicity was tested for all three subgroups (loss of smell, loss of taste, and loss of smell and/or taste) and was highly significant in all three categories with p ≤ 0.0006 (Figs. 3A-C). There were 33 studies available on smell loss in Caucasians and five studies on East Asians. Ethnicity of participants explained a significant amount of heterogeneity in smell loss (Q = 11.9, df = 1, *p* = 0.0006; Fig. 3A). The estimated random prevalence of smell loss was 43.2% [95% CI, 31.9-55.3%] for Caucasians and 15.1% [95% CI, 8.3-25.7%] for East Asians (Fig. 3A). The estimated prevalence of patients with loss of taste was 38.3% [95% CI, 27.0-51.0%] among Caucasians and significantly lower, only 6.4% [95% CI, 5.7-51.0%], among East Asians (Q = 65.87, df = 1, *p* < 0.0001; Fig. 3B). Overall estimates of chemosensory deficits were nearly three times higher for Caucasians than East Asians and also showed high heterogeneity and evidence of publication bias (Figs. 2C and 3C). Differences in chemosensory deficits between East Asians and Caucasians are further illustrated in Figure 4, with the prevalence shown in a world heat map, with the cohort size indicated by the size of the circles, and in the bar graphs in Figure 5.

**Figure 3.**
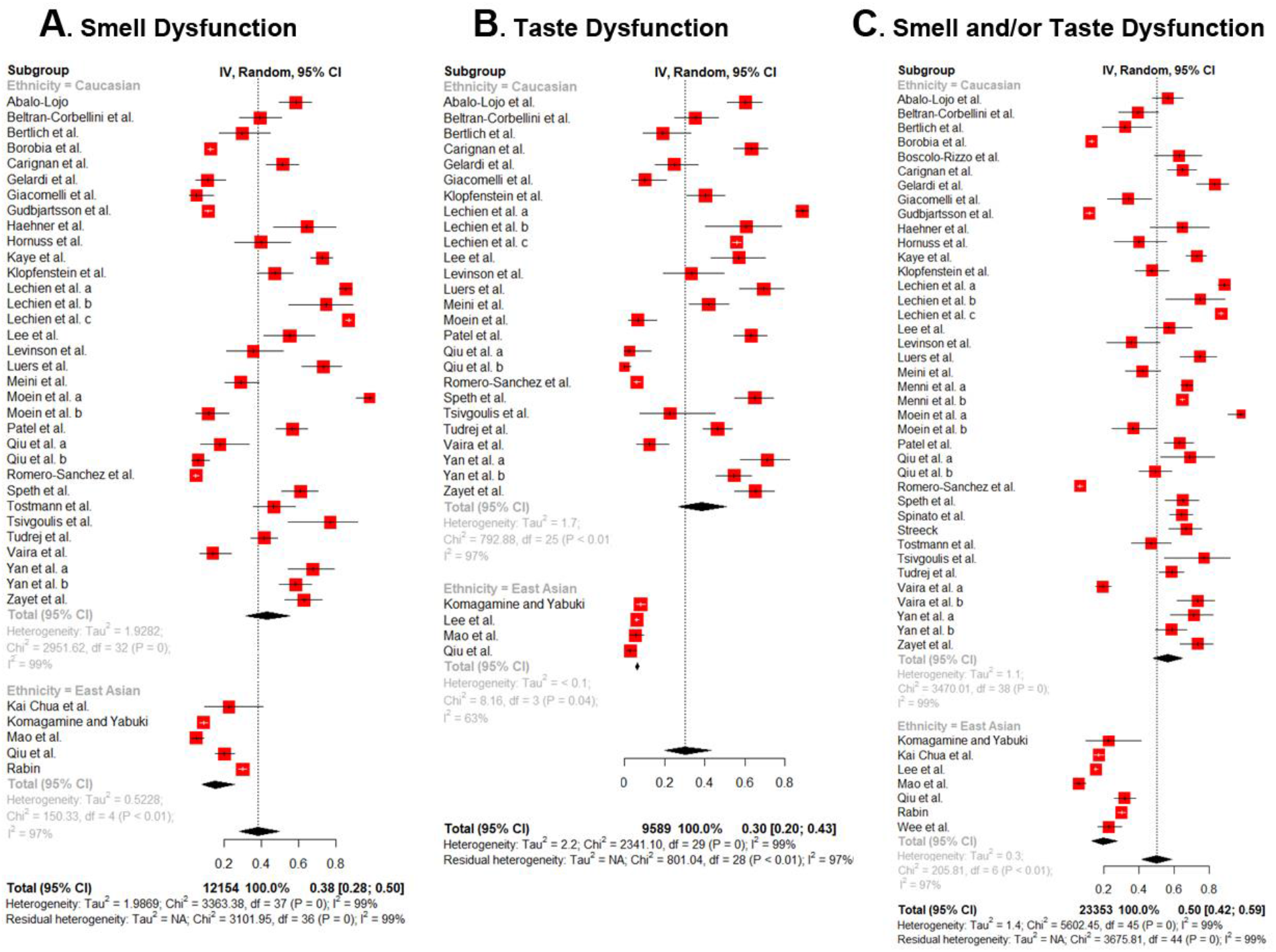
Forest plots of the prevalence of smell dysfunction (A), taste dysfunction (B), and smell and/or taste dysfunction (C) in COVID-19 patients. Estimated random proportions are shown by red boxes with 95% confidence intervals (95% CI) extending as whiskers, the overall estimated random proportion of subgroups is shown in gray, and the results for all studies combined are shown in black. Note the difference between East Asians and Caucasians.

**Figure 4.**
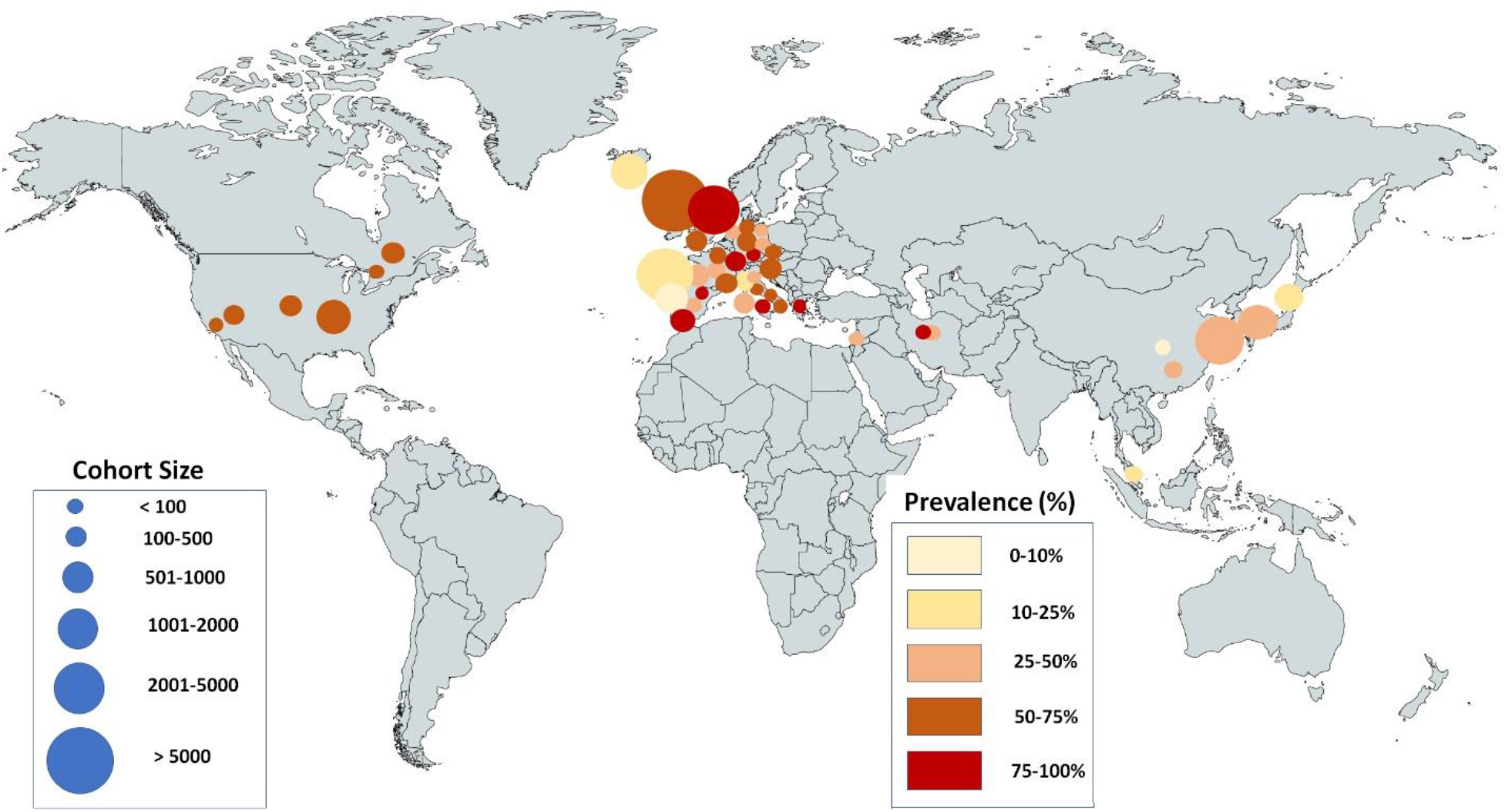
World Map of the Prevalence of any Chemosensory Deficit in COVID-19 Patients

**Figure 5.**
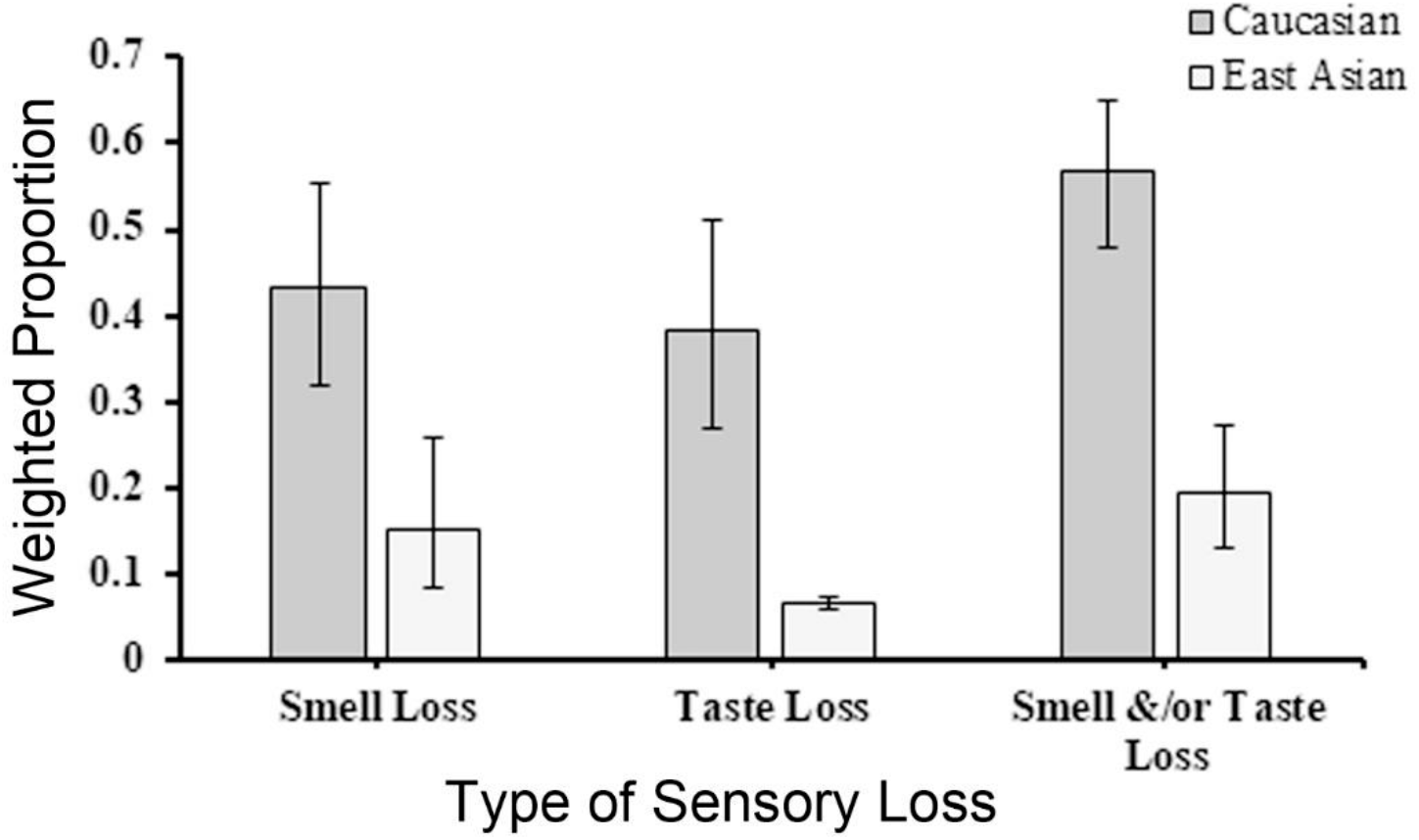
Estimated random prevalence of chemosensory dysfunction in COVID-19 patients, based on ethnicity with 95% confidence intervals from meta-analysis. Note the significant difference between Caucasians and East Asians.

### Disease severity

As a measure of disease severity, we used information about hospitalization rates within cohorts. The weighted regression analyses showed a significant negative influence of the percent of the cohort that was hospitalized during data collection on the proportion of patients with loss of smell, taste, and loss of smell and/or taste (Fig. 6). The beta coefficients for the effect of disease severity on loss of smell (b = -0.0261, *p* = 0.0018) and taste (b = -0.0262, *p* = 0.0035) showed that both were reported less frequently in cohorts as the number of individuals in the cohort who were hospitalized increased. This result was even more highly significant when loss of smell and taste were combined (b = -0.0216, *p* < 0.0001; Fig. 6, Table 2). Accordingly, patients with severe COVID-19 report fewer smell/taste dysfunctions.

**TABLE 2.**
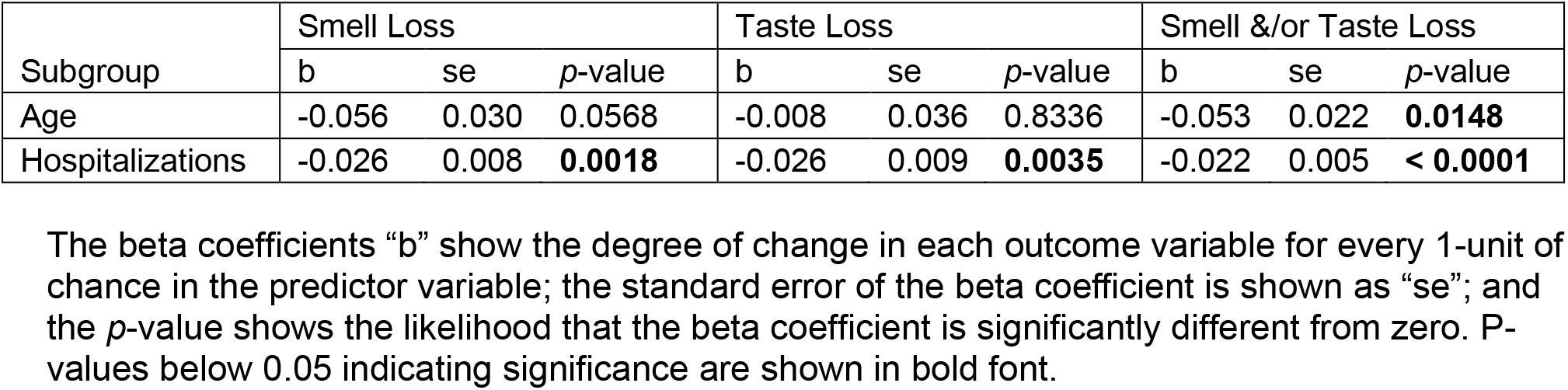
Subgroup test results for continuous variables: age and disease severity (percent of patients hospitalized).

**Figure 6.**
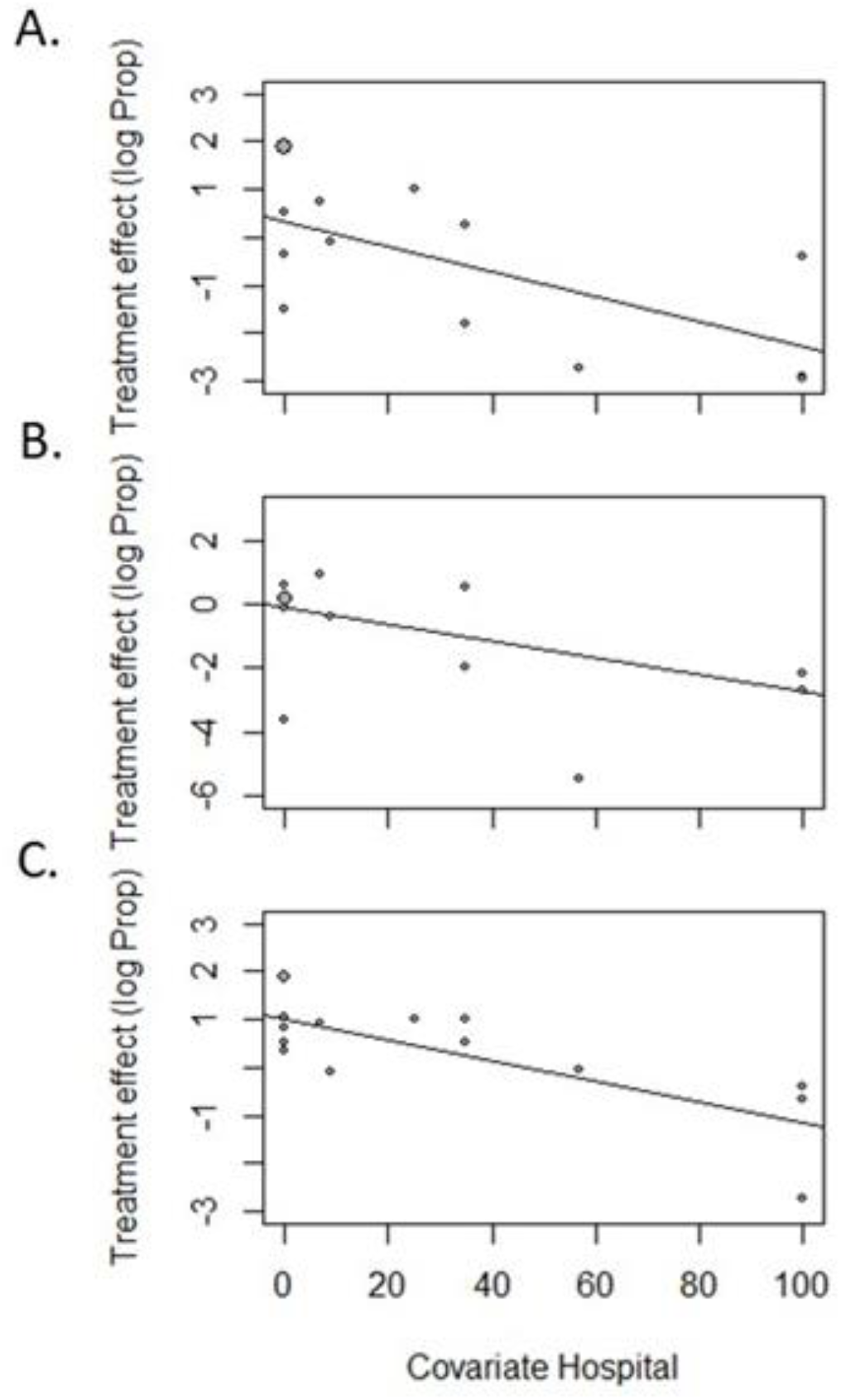
Bubble plots of subgroup tests for relationships between disease severity (the percentage of hospitalized patients) in each cohort and loss of smell (A), taste (B), and loss of smell and/or taste (C). Patients who are hospitalized (have more severe disease) report fewer chemosensory deficits.

### Age

The subgroup tests for the effect of cohort age on smell loss showed some indication of a negative association (b = -0.0562, *p* = 0.0568), while tests for the effect of cohort age on taste showed none (b = -0.0076, *p* = 0.8336; Table 2), and a significant negative association was found when loss of smell and taste were combined (b = - 0.0532, *p* = 0.0148) suggesting that increasing age may result in a lower reporting of loss of sensory deficits in general.

### Methodology

The subgroup test to compare studies that used subjective or objective measures for loss of sensory function was possible only for Caucasians (there were too few studies for East Asians), and it showed no significant differences (in all cases *p* ≥ 0.1564). For loss of smell, studies that used objective measures had an estimated random prevalence of 59.66% [95% CI, 27.16-85.43%] while those that used subjective measures had a prevalence of 42.22% [95%CI, 30.54-54.84%]. For loss of taste, the prevalence among the studies with objective measures was no different than among those with subjective measures (q = 2.01, *p* = 0.1564), with an estimated random prevalence of 25.99% [95% CI, 12.52-46.29%] and 43.12% [CI 95%, 29.99-57.28%], for objective vs. subjective measures. When the two endpoints were combined, the resulting prevalence for objective measures was 69.68% [95%CI, 44.95-86.61%], which again was not significantly different from that of subjective measures (56.85% [95%CI, 48.41-64.91%]; q = 1.00, *p* = 0.3168). Sample sizes were as follows: smell: objective measures = 6, subjective measures = 25; taste: objective measures = 4, subjective measures = 20; smell and/or taste: objective measures = 6, subjective measures = 31.

### Nasal congestion/rhinorrhea

If the anosmia was caused by nasal congestion, as is common in cases of viral infection, then most, if not all COVID-19 patients with anosmia would be expected to have nasal congestion/rhinorrhea. However, our data shows that a weighted mean of 58.6% of COVID-19 patients with anosmia did not have nasal congestion/ obstruction or rhinorrhea, based on n= 17 studies with a total cohort size of 4,121, consistent with the conclusion of a previous report (Lechien et al., 2020d).

### Duration of chemosensory dysfunction

Based on the studies that provided such information, the average duration of smell dysfunction was 9.03 days ± 1.32 (SEM, n= 9 studies with a total cohort number of 2,826), and 12.64 ± 2.51 days for taste dysfunction (n=4 studies with total cohort number of 293). Duration of smell dysfunction appears shorter, but duration of taste dysfunction is based on a small number of studies (n=4).

### Possibility of bias

The majority of the studies are cross-sectional, retrospective observational studies, and therefore, recollection bias may be present. Most studies are similar to those previously graded as “moderate risk of bias” (Tong et al., 2020; see also da Costa et al., 2020). Potential weaknesses are that measures mostly were not validated (Pellegrino et al., 2020), but it has to be considered that data were collected during an unprecedented pandemic when using more time-consuming assessment tools was not possible due to increased risk of virus spreading. The sample size for East Asian studies was small, but with n ≥ 4, it was sufficient for each of the reported comparisons (Fu et al., 2011).

## Discussion

The literature on the prevalence of chemosensory dysfunctions in COVID-19 has been evolving at a rapid pace. In the first two months of the COVID-19 pandemic, such deficits were considered a rare occurrence (Chen et al., 2020; Guan et al., 2020; Mao et al., 2020; Wang et al., 2020; reviewed in da Costa et al., 2020). The first report of smell and taste dysfunction that recognized this condition as a much more prevalent symptom (66.7% of COVID-19 patients) was on March 16^th^, 2020 by a German virologist (Streeck, 2020). The majority of subsequent studies have confirmed such a high prevalence outside of East Asia (Table 1; Fig. 3).

Compared with previous systematic reviews and meta-analyses (da Costa et al., 2020; Passarelli et al., 2020; Pellegrino et al., 2020; Printza and Constantinidis, 2020; Sedaghat et al., 2020; Tong et al., 2020), our review considers a much larger number of studies and cohort numbers. We did not include the study by Bagheri et al. (2020), because subjects in this cohort did not have COVID-19 diagnoses and therefore did not meet the inclusion criteria, although many of the cases likely were related to COVID-19 (Gane et al., 2020). The study by Bagheri et al. (2020) also failed the inclusion criteria of the meta-analysis by Passarelli et al., 2020, but was erroneously included in their analysis.

### Why are chemosensory deficits rare in East Asians with COVID-19 compared to Caucasians?

Some researchers have commented on a possible difference in the frequency of chemosensory deficits between East Asians and Caucasians with COVID-19 (Lechien et al., 2020a; Lavato et al., 2020; Qiu et al., 2020). With the much more extensive datasets considered in our review (16,817 Caucasians and 6,536 East Asians), we show that there indeed is a significant difference in prevalence between these two populations: 3-fold higher for smell, 6-fold higher for taste, and 3-fold higher for smell and/or taste impairment in Caucasians (Fig. 5). Why is there such a difference? It is unlikely that such differences are due to the evolution of the SARS-CoV-2 virus itself. Studies have failed to identify recent virus mutations that increase viral transmission/ infectivity (van Dorp et al., 2020). As pointed out by Lechien et al., 2020a, the differences in the prevalence of chemosensory dysfunction may be due to ethnic differences in the frequency of variants of the ACE2 virus entry protein. Variations in the ACE2 protein can change virus binding by up to 20-fold (Li et al., 2005), and glycosylation sites relevant to the binding may be tissue-specific (Bilinska et al., 2020). ACE2 variants are genetically determined and they are known to differ in frequency between Europeans and East Asians (Benetti et al., 2020; Cao et al., 2020; Strafella et al., 2020; Williams et al., 2020). If Caucasians have more often an ACE2 variant expressed in the olfactory epithelium (presumably in the sustentacular cells of the olfactory epithelium, Bilinska et al., 2020; Butowt and von Bartheld, submitted), then these cells may bind SARS-CoV-2 with higher affinity, resulting in anosmia, whereas East Asians may have less of these ACE2 variants, and therefore will more rarely have anosmia as part of the COVID-19 symptoms.

Since the nasal epithelium has a larger viral load than respiratory epithelium (Zou et al., 2020) – and the nasal epithelium has increased expression of entry proteins for the virus (Bilinska et al., 2020) – this ethnic difference has potentially far-reaching implications for infectivity, spread of the virus (frequency of asymptomatic super-spreaders, Oran and Topol, 2020), and therefore for successful management of the pandemic. The frequency of ACE2 variants may make it more difficult in some ethnicities to control the pandemic, and easier in other ethnicities. The presence of different ACE2 variants in the nose may, in part, explain the more rapid spread of COVID-19 in Caucasians and Hispanics, as compared to East Asians, in addition to the well-known cultural differences in strategies of containment and attitudes about social distancing and the use of protective measures such as face masks.

### Technical aspects and confounding variables

#### Methodology

Most studies rely on the subject telling the researcher about their subjective impressions. A relatively small number of studies (7/42) used objective tests to assess chemosensory dysfunction (Moein et al., 2020; Vaira et al., 2020c; Qiu et al., 2020; Bertlich et al., 2020; Hornuss et al., 2020; Lechien et al., 2020c; Tsivgoulis et al., 2020). When smell and taste was objectively tested, the percentage of subjects with dysfunction increased in some of those studies (Moein et al., 2020; Pellegrino et al., 2020), but in one of these studies, 38% of subjective olfactory loss could not be objectively confirmed (Lechien et al., 2020c). In our analysis, the difference in prevalence between studies with objective vs subjective measures did not reach significance, however, the number of studies using objective measures was small. Self-reporting of anosmia is thought to be relatively accurate (90%, Wehling et al., 2011), so using subjective recall to obtain data on chemosensory deficits appears to be a valid approach, and, in many instances, it is the only feasible way of data collection during a raging pandemic.

#### Olfaction vs. taste

Some of the studies reporting on smell and taste impairment did not examine taste dysfunction separately from smell dysfunction, but rather asked patients about “smell and/or taste dysfunction.” The pooled prevalence of chemosensory dysfunction that we report is likely an underestimate, because many studies reported only how many patients had smell deficits and how many had taste deficits, but they did not report on the potential overlap (there were patients who had both types of chemosensory dysfunction in at least 19/46 cohorts). Those cases were listed in our review conservatively, meaning that we did not simply add all cases with smell dysfunction to those with taste dysfunction, because we know that there is overlap in a substantial fraction of patients (Table 1). Nevertheless, there is no doubt that a large fraction of COVID-19 patients (including otherwise asymptomatic carriers) have chemosensory deficits. Olfaction is used for tasting food (culinary experience) and it can be difficult to subjectively separate the two modalities (Pellegrino et al., 2020). Since most studies asked about *changes* to chemosensory perception, subjects with pre-existing loss of smell or taste would not have been included and would not have given false positives; some studies actively excluded patients with a history of pre-existing anosmia or ageusia.

#### Age

Previous investigators have noted that smell and taste dysfunctions appeared to be more frequent in the younger age groups of COVID-19 patients (e.g., Giacomelli et al., 2020). Our results are consistent with this finding, although significance was reached only for the category “smell and/or taste.” Reduction of smell with age is a well-known phenomenon (Doty and Kamath, 2014), but the sudden loss of function coincident with COVID-19 would still be noticeable in the older population.

#### Duration

Our pooled analysis, based on 9 studies, revealed that the mean duration of the anosmia is 9 days. This relatively short time has implications for the pathogenetic mechanism: It seems too short for a functional recovery if such a recovery involved death and regeneration of olfactory neurons, since their replacement by stem cells alone takes 9-10 days (Schwob et al., 1995; Schwob, 2002). Alternative mechanisms, not requiring neuron death, that may explain the transient anosmia include a support-cell mediated dysfunction of the olfactory epithelium (Heydel et al., 2013) or a virus-induced short-lasting immune response (Butowt and Bilinska, 2020), although the extent of inflammation in the olfactory epithelium in response to SARS-CoV-2 is still unclear (Vaira et al., 2020b; Torabi et al., 2020).

## Methods

We followed the PRISMA guidelines for systematic searches and meta-analyses (Moher et al., 2009). We searched the COVID-19 Portfolio of the National Institutes of Health (https://icite.od.nih.gov/covid19/search/) with the key words „anosmia”, “smell,” or “taste” on and before June 10, 2020, resulting in 1,103 records, including preprints posted prior to peer review. We also examined and included any relevant references within, and citations of, screened records. Inclusion criteria were that the paper was a novel report of the prevalence of smell and/or taste impairment in patients verified to have COVID-19. We accepted all studies that reported original and quantitative data on prevalence of chemosensory deficits in human subjects, either obtained by questioning the subjects, by chart review, or by objective chemosensory testing. We excluded from our quantitative analysis case reports, reports that did not provide exact quantitative information, reviews only, and reports of cohorts in which a COVID-19 diagnosis was not confirmed clinically or by lab tests. We also excluded studies that targeted *any* patients with chemosensory deficits, regardless of cause, because they would fail to provide a true prevalence among COVID-19 (Bagheri et al., 2020; Parma et al., 2020). We kept data on the two senses, olfaction and taste, separate, when the study reported them separately. The most common way of reporting in studies was “smell deficit,” “taste deficit,” or “smell and/or taste deficit,” and those data were separately compiled and compared. For this reason, the cohort number for olfactory deficits and gustatory deficits differs from that of the combined (smell and/or taste) category.

A pooled analysis was performed for prevalence, and significance and confidence intervals were calculated in the software “R”. We used R Studio, version 1.2.1335, for statistical analyses (R Foundation for Statistical Computing, Vienna, Austria). To calculate estimates of pooled prevalence and 95% confidence intervals, we used the R-meta package, version 4.9-5, and the metaprop function. We used random effects models with the inverse variance method for pooling and the logit transformation for proportions (DerSimonian and Laird, 1986). For ease of interpretation, we back transformed and rescaled proportions to events per 100 observations. Analysis of the heterogeneity across studies was done using the Maximum-likelihood estimator, Higgin’s I^2^ and Cochran’s Q method (DerSimonian and Laird, 1986; Higgins and Thompson, 2002). Publication bias was assessed by visual inspection of funnel plots (Egger et al., 1997). In all cases, significance was defined at α = 0.05. Subgroup analysis was conducted by ethnicity, age, hospitalization rate, and methodology. Ethnicity was coded as a categorical variable with two levels: Caucasian and East Asian, because of suspected heterogeneity (Lechien et al., 2020a; Lavato et al., 2020; Qiu et al., 2020) and because these two ethnicities are the only ones for which such data are currently available. All other subgroup tests used continuous variables and the metareg function to adjust the overall meta-analysis for the subgroup. The subgroup age was a created variable that uses the center of the sample, either the mean or the median, to mark the center of the age distribution. Hospitalization rate was the percentage of subjects in the sample that were hospitalized for COVID-19.

## Data Availability

All data referred to in the manuscript are available.

## Author Contributions

Christopher von Bartheld and Rafal Butowt designed the study and wrote the manuscript, Molly Hagen prepared the meta-analysis, and all authors edited and approved the final version of the manuscript.

